# How high and long will the COVID-19 wave be? A data-driven approach to model and predict the COVID-19 epidemic and the required capacity for the German health system

**DOI:** 10.1101/2020.04.14.20064790

**Authors:** Thomas Klabunde, Clemens Giegerich

## Abstract

**Background and objective:** In March 2020 the SARS-CoV-2 outbreak has been declared as global pandemic. Most countries have implemented numerous “social distancing” measures in order to limit its transmission and control the outbreak. This study aims to describe the impact of these control measures on the spread of the disease for Italy and Germany, forecast the epidemic trend of COVID-19 in both countries and estimate the medical capacity requirements in terms of hospital beds and intensive care units (ICUs) for optimal clinical treatment of severe and critical COVID-19 patients, for the Germany health system.

**Methods:** We used an exponential decline function to model the trajectory of the daily growth rate of infections in Italy and Germany. A linear regression of the logarithmic growth rate functions of different stages allowed to describe the impact of the “social distancing” measures leading to a faster decline of the growth rate in both countries. We used the linear model to predict the number of diagnosed and fatal COVID-19 cases from April 10^th^ until May 31^st^. For Germany we estimated the required daily number of hospital beds and intensive care units (ICU) using clinical observations on the average lengths of a hospital stay for the severe and critical COVID-19 patients.

**Results:** Analyzing the data from Germany and Italy allowed us to identify changes in the trajectory of the growth rate of infection most likely resulted from the various “social distancing” measures implemented. In Italy a stronger decline in the growth rate was observed around the week of March 17^th^, whereas for Germany the stronger decline occurred approximately a week later (the week of March 23^rd^). Under the assumption that the impact of the measures will last, the total size of the outbreak can be estimated to 155,000 cases in Germany (range 140,000-180,000) and to 185,000 cases in Italy (range 175,000-200,000). For Germany the total number of deaths until May 31^st^ is calculated to 3,850 (range 3,500-4,450). Based on the projected number of new COVID-19 cases we expect that the hospital capacity requirements for severe and critical cases in Germany will decline from the 2^nd^ week of April onwards from 13,500 to ∼2500 hospital beds (range 1500-4300) and from 2500 to ∼500 ICU beds in early May (range 300-800).

**Conclusions:** The modeling effort presented here provides a valuable framework to capture the impact of the “social distancing” measures on the COVID-19 epidemic in European countries and to forecast the future trend of daily COVID-19 cases. It provides a tool for medical authorities in Germany and other countries to help inform the required hospital capacity of the health care system. Germany appears to be in the middle of the (first) COVID-19 outbreak wave and the German health system is well prepared to handle it with the available capacities.

## Introduction

On March 11^th^ 2020 the world health organization (WHO) declared the COVID-19 outbreak as pandemic. The steep increase of COVID-19 cases in China, Iran, Italy, France, Germany and other European countries resulted in to tens of thousands of severe cases requiring hospitalization as well as in thousands of critical cases due to progression to life-threatening respiratory distress that require intensive care unit (ICU) admission (Fig. 1). The dramatic rate of cases that progresses has created a significant strain in the national health systems of these countries and has led to a significant shortage of ICUs equipped with extra-corporeal membrane oxygenation (ECMO) to save the lives of very critical COVID-19 cases.

**Figure 1.**
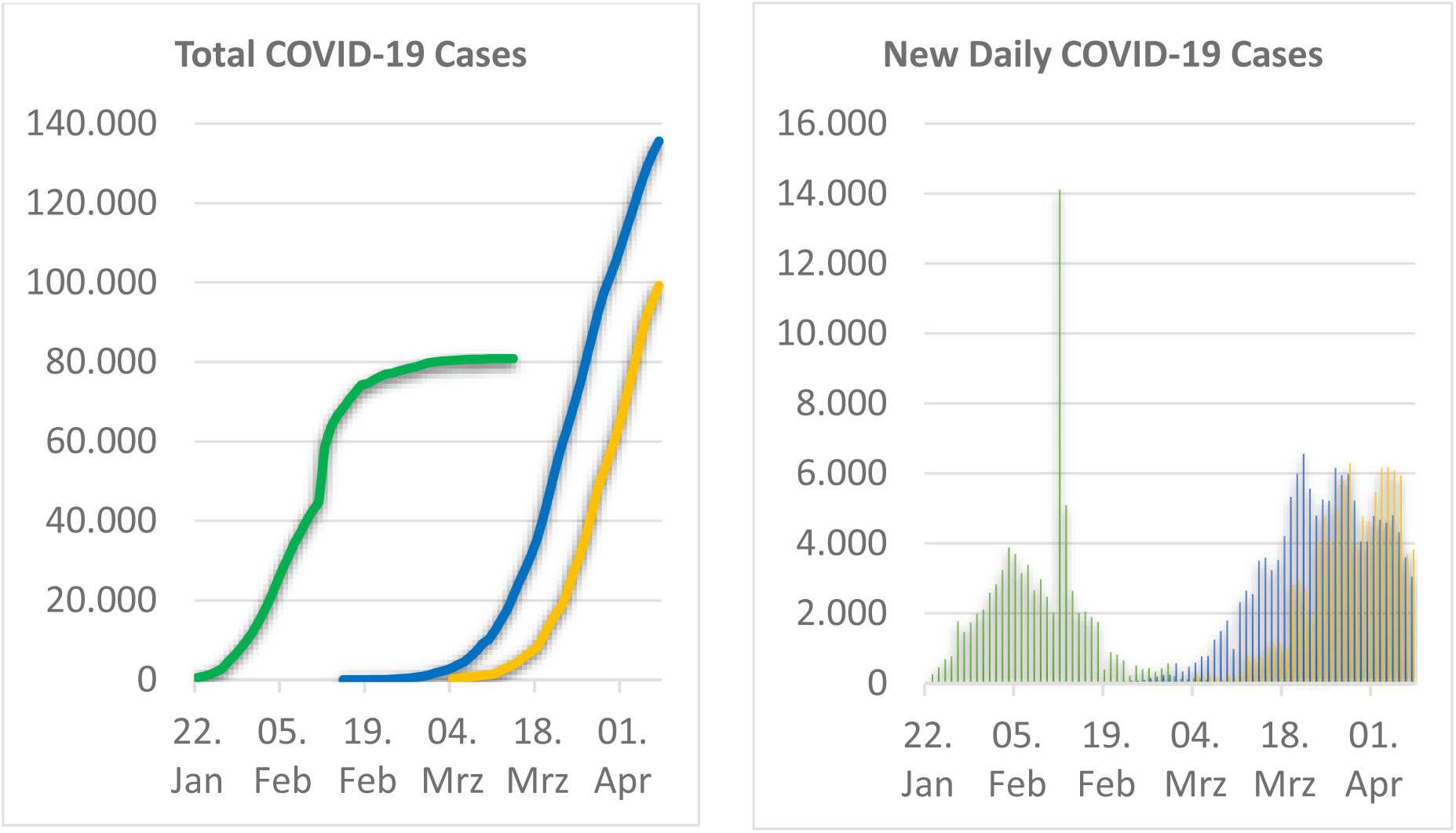
Total cumulated (left) and new daily (right) COVID-19 cases from Jan 22^nd^ to April 7^th^ 2020 in China (green), Italy (blue) and Germany (orange). Data source: China and Italy from John Hopkins University (JHU), Germany from Robert Koch Institut (RKI) for Germany.

Since early March several European governments have implemented numerous control measures to reduce the transmission of the disease and decrease the number of new daily cases of COVID-19 so that fewer patients need to seek treatment at any given time and avoid overwhelming hospital capacity, commonly referred as “flattening the curve”. The strategy to achieve of the flattening of the curve of COVID-19 cases in Europe has consisted of (1) early detection of new cases and primary contacts followed by self-quarantine or hospitalization, (2) promoting the adherence to hygiene standards like washing hands and (3) restricting contacts between potentially infectious and susceptible parts of the population by “social distancing”. In Germany these “social distancing” measures have evolved over time from banning gatherings of more than 1000 participants (issued on March 9^th^), university and school closures (issued on March 16^th^), to prohibiting gatherings of more than two people (issued on March 23^rd^). These restrictions have put a significant burden on German society and have reduced the quality of life for most. There has been a mounting interest on how long these measures and restrictions will last and when can they be lifted safely. On the other hand, medical professionals are expecting a surge of COVID-19 patients reaching the hospitals soon and wonder when and how hard it will hit them.

Germany has significantly expanded its hospital capacity by increasing the number of ICU beds and ECMO units in order to prepare for the expected wave of COVID-19 patients and to prevent overwhelming the health system. A registry has been formed to track all occupied and readily available intensive care beds in Germany to ensure that possible regional peaks of critical COVID-19 cases can be treated by transferring the influx of patients to nearby health systems with availability [1]. As of April 12^th^ this registry covers approximately 19,700 ICU beds with 11,376 occupied beds and 8325 available beds. This indicates that there is available capacity for any upcoming influx of new critical and severe cases.

Over the past weeks mathematical modeling of the COVID-19 pandemic – especially SIR (susceptible, infectious, recovered) models – has provided important insights to better understand the spread of virus SARS-CoV-2 and to evaluate how various potential scenarios of control measures could impact the dynamics of the COVID-19 out-break [2,3]. These simulation studies have informed decision making for national governments on how to react to this coming COVID-19 epidemic. Simulated estimates of the size of the epidemic in Germany if no control measures were implemented [4] and have provided a strong rationale for the closures of school and universities in the UK [5]. Other simulations have shown that a contact-tracing App that builds a memory of proximity contacts and immediately notifies contacts of positive cases – and potentially asks for self-isolation – might significantly reduce the transfection rate especially by pre-symptomatic COVID-19 cases thus reducing the spread of the virus [6].

Due to the underlying mechanistic model structure of SIR-based models that include representations of mobility information of society it is possible to explore multiple control measure scenarios and thus estimate the impact of certain measures at a very early stage of the epidemic. Usually, an epidemic follows an exponential growth at an early stage, peaks and then at the inflection point of the total case curves the growth rate declines again. This decline of the growth rate is either achieved by reaching herd immunity or by implementation of measures to hinder the transmission of the virus. As soon as the impact of these measures become apparent in the decline of the daily growth rate, empirical or so-called top-down methods can be applied to model the data (e.g. logistic growth model, Gompertz model, exponential growth rate decline model). These top-down modeling methods can then provide insights regarding the stage of the epidemic, allow the prediction of future trends of the outbreak and estimate its final size [7-9].

Since early April 2020 the numbers of daily new COVID-19 cases in Germany and Italy appear to be either constant with similar numbers of new infections each day or declining (see Figure 1), indicating that the epidemic has moved from the exponential growth phase into a linear growth phase in these countries. With this new data becoming available, we have used an exponential decline of the growth rate model to analyze the trajectory of the daily growth rate [10]. Thus, we have captured and quantified the dynamics of the epidemic in these countries and described the impact on the growth rate measures have had taken. Using this approach, we have captured a specific date for both countries – two to three weeks after the start of these measurements – when the spread of the epidemic significantly slowed down. In a second step we have used the model to forecast the dynamics of the epidemic in Italy and Germany and to estimate the number of daily and cumulative total COVID-19 cases. This allowed us to address the question of how long the epidemic will last in both countries. In a third step we have used these forecasts of diagnosed COVID-19 cases to estimate the number of severe and critical cases for each day of April and May to provide an estimate for the daily required capacity of hospital beds, in particular ICU beds with ECMO equipment to treat the predicted severe and critical COVID-19 cases in Germany.

## Material and Methods

### Modeling COVID-19 epidemic in Germany and Italy using exponential decline growth rate model

We have used an *exponential decline of growth rate* model to analyze the trajectory of the daily growth rate in Germany and Italy. We also evaluated two well established growth models, the logistic function and the Gompertz function, but decided in favor of the exponential decline model as the model structure allows to easily capturing differences in the growth curve that may result from the social distancing measures. The exponential growth rate decline model defines the growth rate as ratio of new cases today versus total cases yesterday as shown in equation (1) and assumes the growth rate is following an exponential decline function given in equation (2). The parameter τ characterizes how fast the growth rate is decreasing. High values of τ indicate a slow decline of the growth rate, whereas low values indicate that the growth rate is declining fast and that the ratio of new cases versus total cases will reach zero more rapidly. With the given model a logarithmic plot of the daily growth rate trajectory thus provided a straight line and allowed us to derive τ from the slope by linear regression.

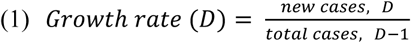

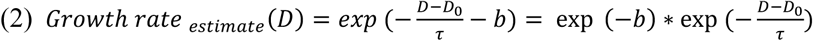

Assuming the measures have an impact on the decline of the growth rate, we would expect a discontinuity in the logarithmic plot of the data. For each country we obtained two fits with different values for τ, capturing the dynamics before and after the impact of the implemented control measures became visible.

For Germany data was obtained from the website of the Robert Koch Institute (RKI) [11]. For Italy the data was taken from the *Coronavirus COVID-19 Global Cases* published by the Center for Systems Science Engineering (CSSE) at Johns Hopkins University (JHU) [12]. For Germany and Italy, we used the reported data up to April 9^th^ for modeling and parameter estimation.

### Simulations of the dynamic and size of COVID-19 epidemic in Germany and Italy

As a second step we used the *exponential decline of growth rate* model to predict the cumulative diagnosed COVID-19 and the daily new cases for Germany and Italy beyond April 9^th^. As described before a linear regression of the logarithmic plot of the growth rate data between March 23^rd^ and April 9^th^ for Germany and between March 17^th^ and April 9^th^ for Italy, respectively, allowed us to model the growth rate using equation (2). Here we could identify the values for τ and the intercept as well as their standard deviation and confidence intervals. This allowed us to estimate the expected growth rate for each day beyond April 10^th^, as well as an upper and lower limit of the growth rate based on the 95% confidence interval of the linear model. For the prediction of the number of total cases for April 10^th^ onwards by equation (3) we calculated the expected number of total COVID-19 cases using the calculated growth rate value and its upper and lower limit. This allowed us to project the uncertainty in the parameter identification from the logistic regression model into a confidence interval for the predicted COVID-19 cases.

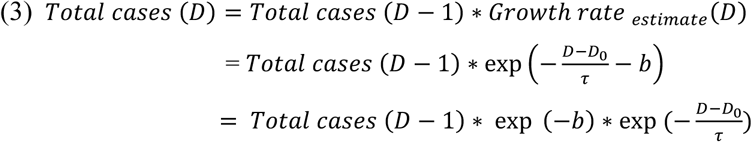

### Simulations of daily required capacity of hospital beds and intensive care units for treatment of severe and critical COVID-19 cases in Germany

We have used the predicted number of COVID-19 cases (including the confidence intervals of these predictions) in the remaining of April and May to estimate the daily required capacity of hospital beds, in particular ICU beds with ECMO equipment to treat the predicted severe and critical COVID-19 cases in Germany. We used assumptions adapted from Thomas-Rüddel et al. [13] on the fraction of diagnosed COVID-19 cases requiring hospitalization and the subset of cases that progress to intensive care along with the average length of each disease state of an COVID-19 patient (summarized in Figure 2). After first symptoms appear on day 0 for an average COVID-19 patient, we assume ∼7 days to reported diagnosis (labeled with D in Figure 2) [14]. For severe cases we assume ∼7 days from first symptoms to hospitalization [15], for critical cases ∼8 days from first symptoms to ICU and for fatal cases ∼14 days from first symptoms to death (labeled with M in Figure 2) [16]. Thus we expect a time period of ∼7 days between reported diagnosis and death. We assume an average duration of hospital stay for severe and critical cases of 12 days [17], an average duration of a stay in intensive care unit of 6 days for fatal critical cases and of 12 days for critical cases that recover [17]. We assume that ∼20 % of COVID-19 cases are severe and require hospitalization, ∼25% of the severe cases developed into critical cases requiring intensive care and ∼50% of the ICU patients have a fatal outcome [17].

**Figure 2.**
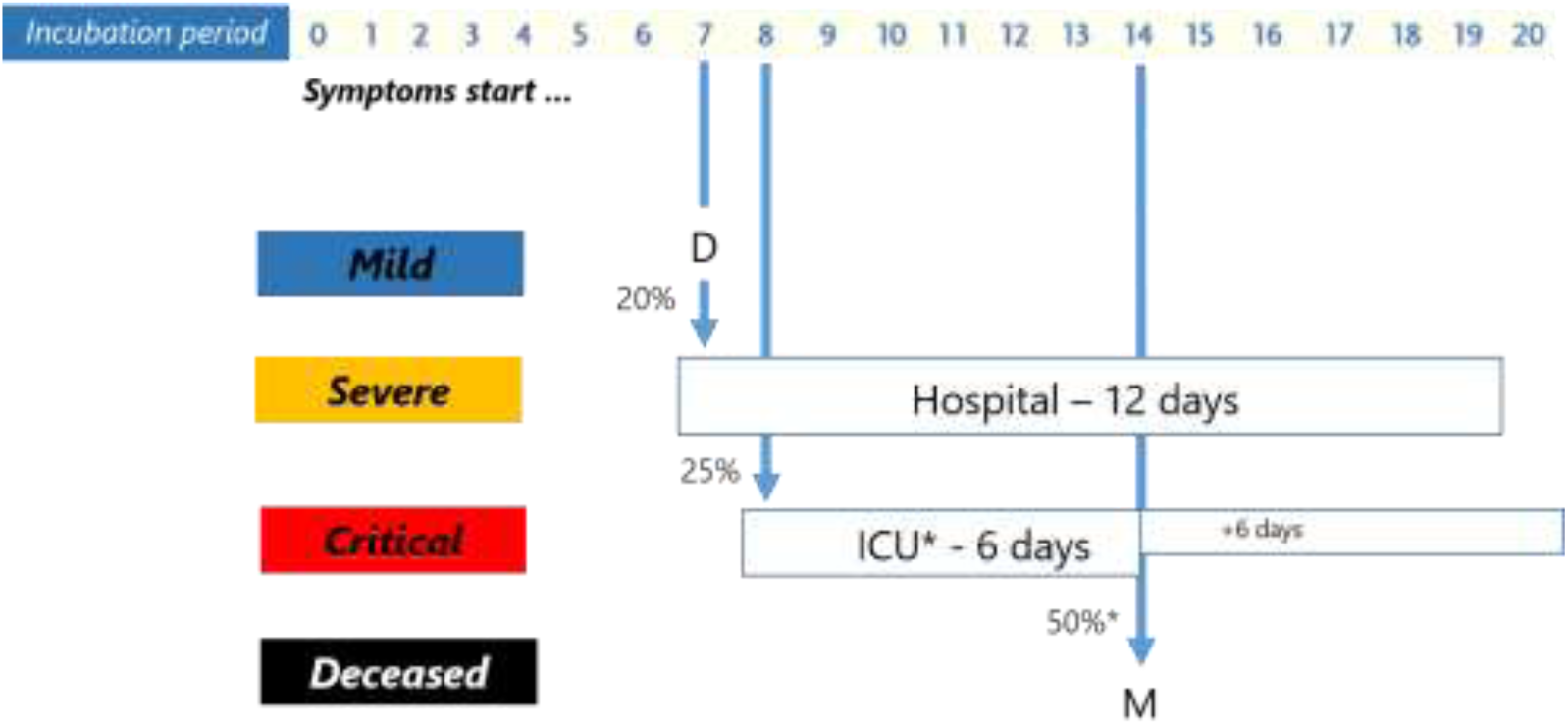
Assumptions used for predictions on the fraction of diagnosed COVID-19 cases requiring hospitalization and/or intensive care and on the average length of each disease state of an COVID-19 patient. D labels the diagnosed COVID-19 patient, M labels a fatal COVID-19 case.

## Results

### Modeling COVID-19 epidemic in Germany and Italy using exponential decline growth rate model

We have used an exponential decline of the growth rate model – with the growth rate being defined by daily new cases today versus daily total cases yesterday – to analyze the trajectory of the daily growth rate in Germany and Italy. Figure 3 shows the plot of the daily growth rate – using a logarithmic function for linearization – from March 1^st^ to April 9^th^ in Italy and Germany. By fitting the data using linear regression, we obtained two fits of the data that are shown in blue and orange.

**Figure 3.**
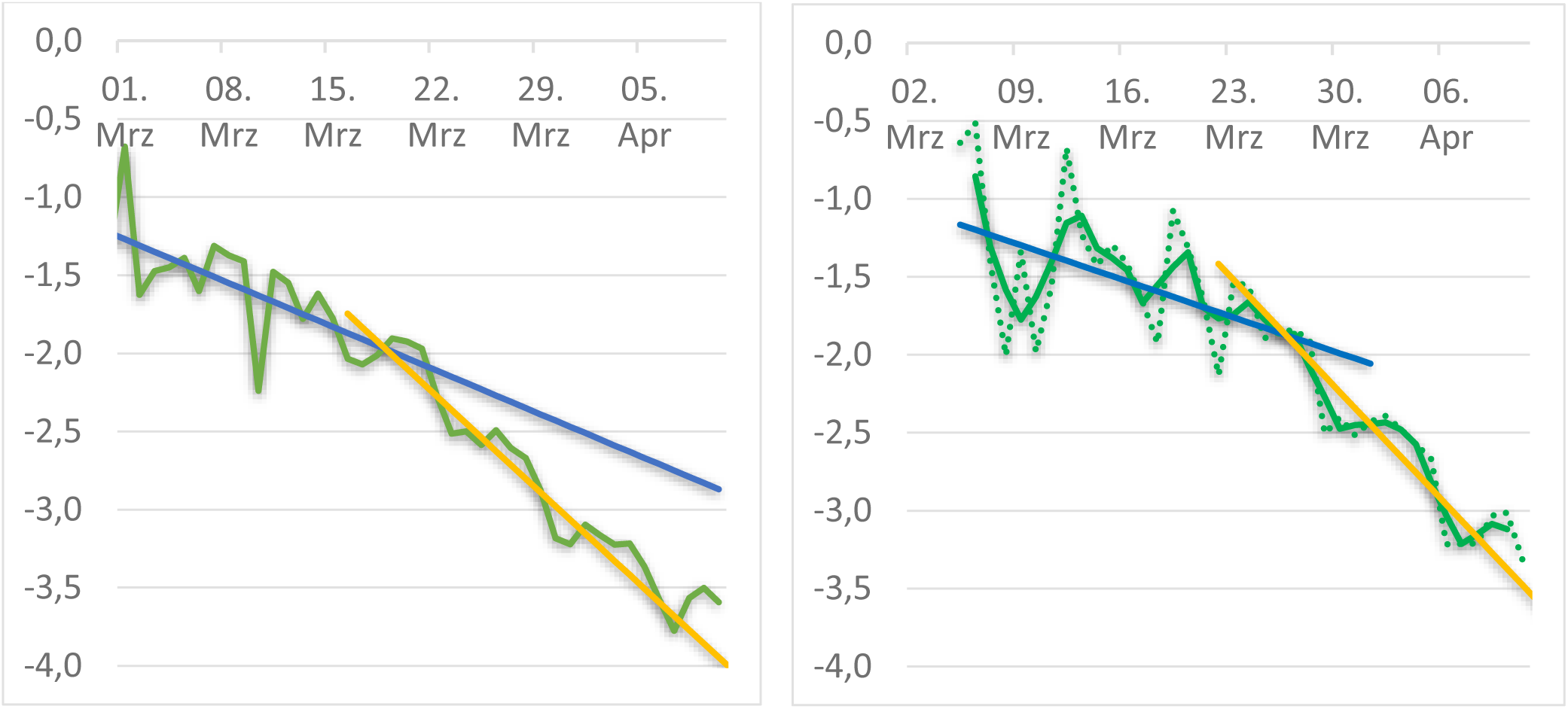
Daily growth rate (new cases at given day / total cases at previous day) of COVID-19 cases for Italy (left) and Germany (right) shown as logarithmic plot. Two linear regression fits (blue and orange) with different values for τ are shown for both countries matching the data for a time period before and after the control measures. Data is shown as green line, data for Germany has been smoothened (3-day average is shown as solid line, original data as dotted line).

For Italy (Figure 3, left panel) initially the growth rate declines with a value for τ of 21.2 days. The impact of the control measures become visible approximately around March 17^th^, two to three weeks after the travel restrictions have been issued on March 1^st^. After March 17^th^ the growth rate shows a faster decline with a value for τ of 12.4 +/-0.6 days (-1/τ = 0.081 +/-0.004).

For Germany (Figure 3, right panel) the growth rate initially declines slowly with a value for τ of 30.5 days. Around March 23^rd^ and thus approximately 2 weeks after first control measures have been issued in Germany (banning of gatherings of more than 1000 participants on March 9^th^), the growth rate shows a much faster decline with a value for τ of 10.3 +/-0.9 days (-1/ τ = 0.097 +/-0.008).

### Simulations of the dynamics and size of COVID-19 epidemic in Germany and Italy

With the value for τ being identified we predicted the trend for new daily cases and accumulated total cases for Germany and Italy assuming that the same control measures remain in place until the last day of May. We projected beyond April 10^th^ using a value for τ of 10.3 and 12.4 days for Germany and Italy, respectively, that has been determined from the regression analysis using data from the day the impact of the measures have become visible until April 9^th^. Figure 4 shows the observed data in blue for the total and daily new COVID-19 cases and the projected cases in the same graph in red. For the projections of the total COVID-19 cases an estimate of the total case range (upper and lower bound) is given that results from the uncertainty in the parameter identification from the linear regression.

**Figure 4:**
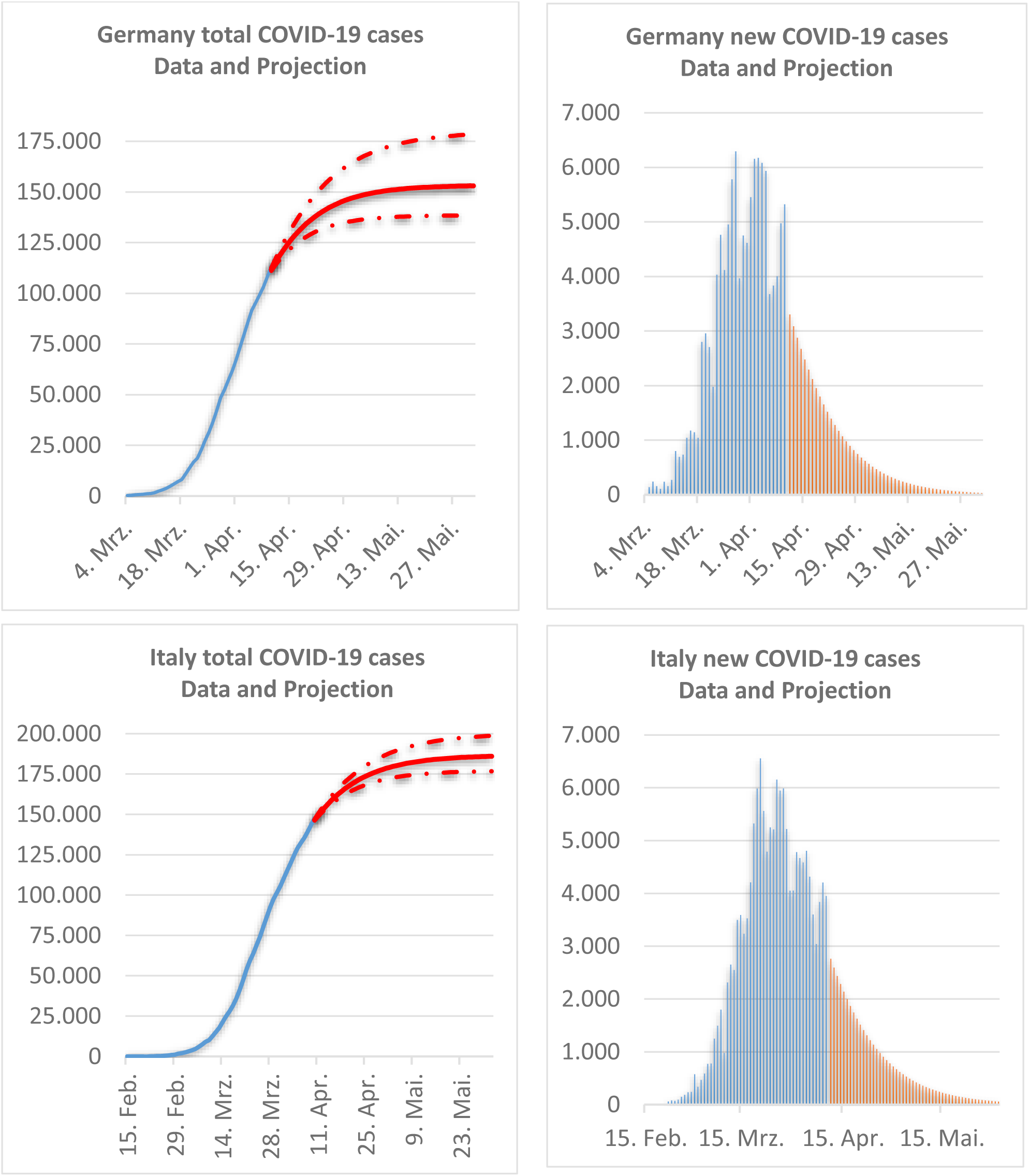
Data and model-based predictions of total (left) and new (right) COVID-19 cases for Germany (top) and Italy (bottom). Data is shown in blue, model-based predictions are shown in red. Projections start on April 10^th^ and use a value for τ to describe the decline of the growth rate that has been derived from data at the day when the impact of measures on the growth rate has become visible to April 9^th^. For the model-based predictions of the total COVID-19 cases an upper and lower bound is given reflecting the uncertainty in the parameter identification (based on the linear model 95% confidence interval).

Based on these projections the total number of cases in Germany by end of May can be predicted to be 140,000 to 180,000 with a peak in the cases around April 1^st^. For Italy the estimated total size of the epidemic is predicted to be 175,000 to 200,000 cases. Evidently these projections assume that the decline of the daily number of new cases continues to follow the exponential trend due to successful control measures taken by both countries.

### Simulating the required daily capacity of hospital beds and intensive care units for treatment of severe and critical COVID-19 cases in Germany

How high will be the COVID-19 patient wave for the German health system? When will it hit? How many people will die? We have used the prediction of the newly diagnosed COVID-19 cases for Germany to estimate the daily capacity requirements for hospital beds and ICUs as well as daily death rates for April and May in Germany.

Figure 5 reveals the daily COVID-19 deaths in Germany: observed data is shown in blue, calculated numbers are shown in red. Calculations are based on the assumption that on average 2.5% of the diagnosed cases will have a fatal outcome 7 days after reported diagnosis (see methods and Figure 2). They use the observed number of diagnosed COVID-19 cases as input before April 10^th^ and the number of projected COVID-19 cases after April 10^th^. The total number of deaths until May 31^st^ is estimated calculated to 3850 (range 3508 to 4450 using the growth rate confidence interval of 95% for the expected diagnosed patients beyond April 10^th^).

**Figure 5.**
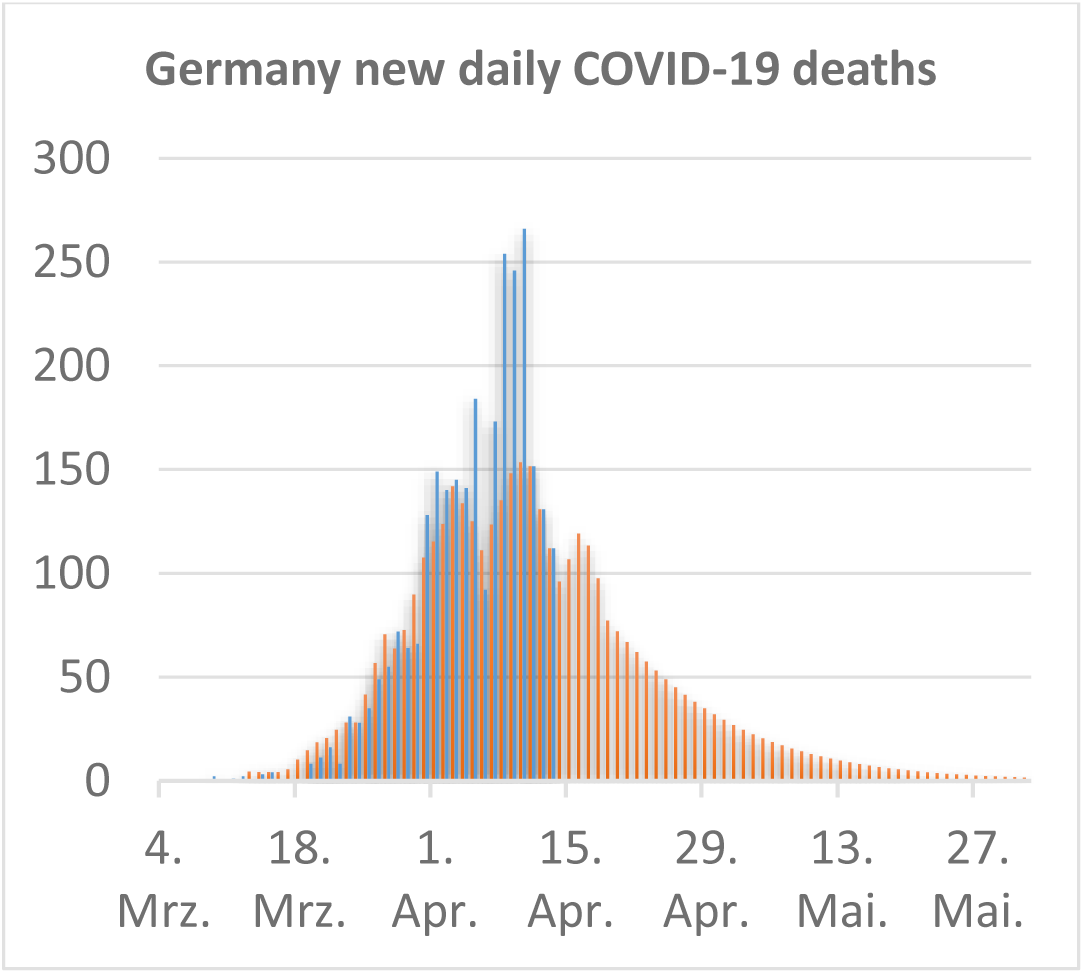
Data and model-based predictions of fatal COVID-19 cases for Germany. Data is shown in blue, calculated values in red (assuming a death rate of 2.5% from diagnosed COVID-19 cases 7 days before).

Next, Figure 6 provides an estimate for every day in April or May for the required capacity of hospital beds, in particular ICU beds with ECMO equipment, to adequately treat the predicted severe and critical COVID-19 cases in Germany. We made assumptions on the fraction of mild, severe, critical or fatal COVID-19 cases and on the average length of the required hospitalization and/or intensive care for each disease state (see methods and Figure 2). We used the observed number of diagnosed COVID-19 cases as input before April 10^th^ and the number of projected COVID-19 cases after April 10^th^. For the projections beyond April 10^th^ a confidence interval is given that results from the uncertainty in the parameter identification from a linear regression. According to the model the number of severe cases requiring hospitalization is expected to peak in the first week of April at ∼13,000. The maximum of ∼2500 critical cases requiring intensive care with ECMO capability is expected at in the first week of April. This is in good agreement with the reported number of COVID-19 cases on ICUs in Germany of 2.405 on April 12^th^ [1, 11]. Based on the expected number of new COVID-19 cases we would expect that the capacity needs for severe and critical cases will decline from the 2^nd^ week of April onwards from ∼13,500 to ∼2500 (range 1500 to 4300) for hospital beds and from ∼2500 to ∼500 (range 300 to 800) for ICU units, respectively.

**Figure 6.**
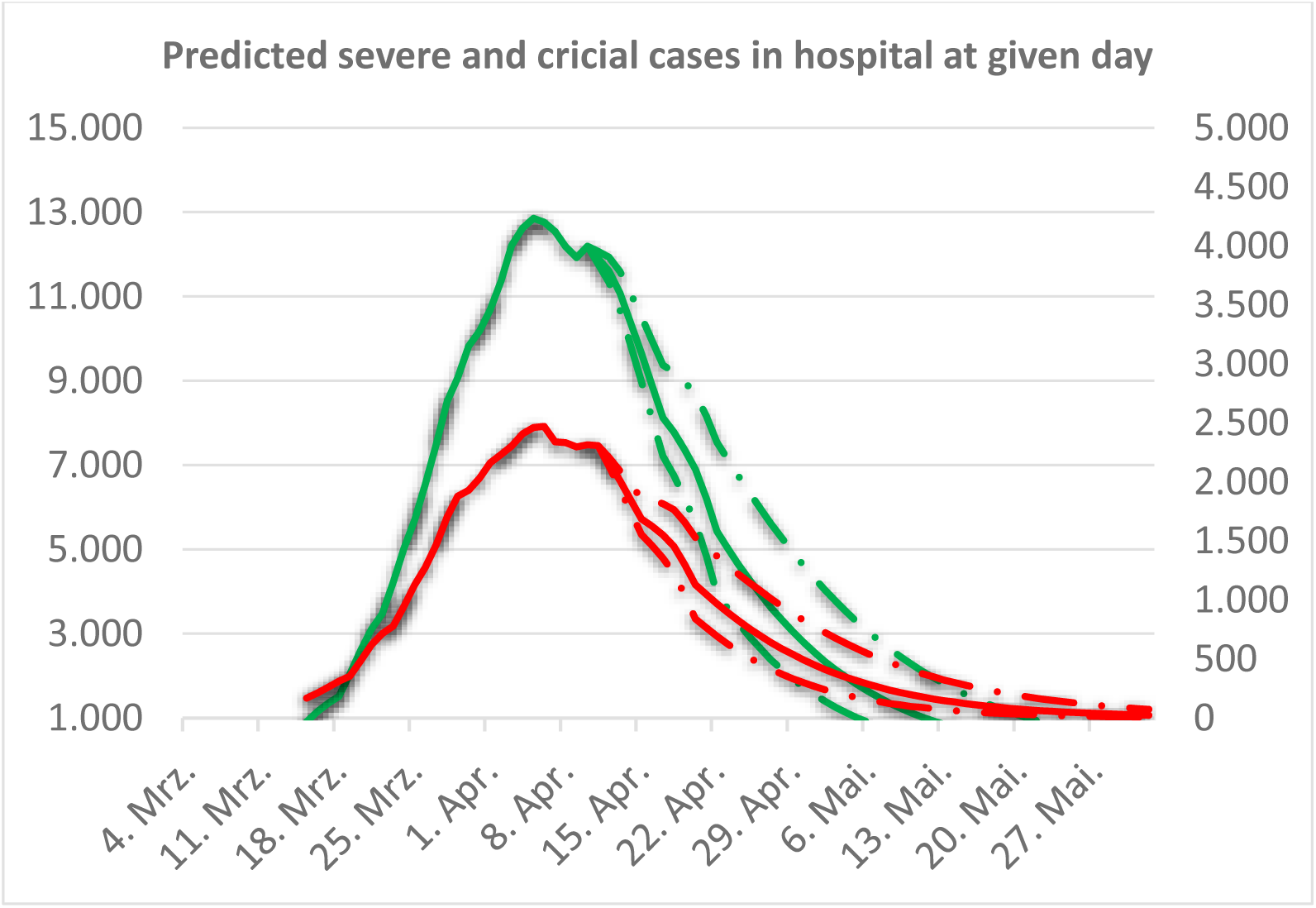
Expected impact of COVID-19 wave on German health system: Daily expected capacity needs of hospital beds (green line) and ICU units (red line) at a given day. For the model-based predictions beyond April 10^th^ of the fraction of total COVID-19 cases an upper and lower bound (red dashed line) is given reflecting the uncertainty in the parameter identification (based on the linear model 95% confidence interval).

## Discussion

In this study we have captured the trajectory of the daily growth rate of COVID-19 cases in Germany and Italy with an exponential decline model. Analyzing the data from Germany and Italy allowed us to identify changes in the trajectory of the growth rate that most likely are the result from the various “social distancing” measures that have been implemented to reduce the transmission of COVID-19. For Italy a stronger decline in the growth rate has been observed starting around the week of March 17^th^, for Germany approximately one week later around March 23^rd^. For Germany this change in the growth rate trajectory could be the result of several measures of “social distancing”, which have been issued since March 9^th^ (banning of gatherings of more than 1000 participants), March 16^th^ (closures of university and school closures) and March 23^rd^ (prohibition of gatherings of more than two people). With the given delay between infection and reported COVID-19 diagnosis of approximately 12 days it can be hypothesized that a significant part of the observed effect on the growth rate can be attributed to the banning of larger gatherings such as soccer matches of national and international leagues.

Beyond the analysis of the given data the data-driven modeling approach presented allowed to describe the observed data with a mathematical model and to provide predictions of new and cumulative COVID-19 cases for both countries. The predictions suggest that the peak of the epidemic in terms of newly diagnosed cases has passed not only in Italy (in late March), but that also in Germany (early April). Under the assumptions that the impact of the control measures on the transmission of the virus will last for the duration of the simulated time period, the total size of the epidemic can be estimated to be 155,000 cases for Germany (range 140,000 to 180,000) and to be 185,000 cases for Italy (range 175,000 to 200,000) for Italy. Evidently these projections assume that the decline of the daily number of new cases will continue to follow the exponential trend due to successful measures taken in both countries.

We have used the predicted daily new diagnosed COVID-19 cases in Germany to estimate the number of daily new severe, critical and fatal COVID-19 cases allowing us to estimate the actual requirements for hospital beds and ICUs at any given day for April and May. Based on the projected number of new COVID-19 cases we would expect that the capacity needs for severe and critical cases will decline from the 2^nd^ week of April onwards from ∼13,500 to ∼2500 hospital beds in early May (range 1500 to 4300) and from ∼2500 to ∼500 ICU beds in early May (range 300 to 800), respectively. The German hospital register tracks all occupied and available intensive care beds in Germany [1]. As of April 12 this register covers approximately 19,700 ICU beds with 11,376 occupied beds and 8,325 available beds. Compared to the expected now decreasing demand for ICU beds this indicates that there is sufficient capacity for the new critical and severe COVID-19 cases. In other words, we are just in with respect to the requirements for the German hospitals middle of the wave -that has been expected to hit and possibly overwhelm the German health system -has been reached and it can be well covered by the available capacities. Our analysis covers Germany as a single entity and does not offer the granularity that may be needed to identify regional short comings of hospital capacities in areas that are more severely effected by the COVID-19 epidemic.

Evidently projections in the future come with certain limitations. First the projection for new COVID-19 cases assume a continuation of (similar) control measures of social distancing having the same impact on the growth rate of new cases. They can only provide a trend, can’t be applied to project further out into the future and obviously are not able to predict the size of a second wave (or whether there would be one at all). In addition, for the calculations used above to estimate the risk for the overwhelming of the German health care system, several assumptions are based on clinical data from the epidemic in China. Evidently due to differences in the age distribution of patients and in the health system between both countries these can only be estimates for the situation in Germany. Nevertheless, it appears that several of these assumptions translate into projections of death rate and capacity needs for ICU beds that are in-line with the observed clinical findings in Germany. We estimate the total rate of fatalities per diagnosed COVID-19 case to be 2.5% for Germany, resulting from the assumption that 20% of the diagnosed cases are severe, 25% of the severe cases get critical and 50% of the critical cases are fatal. On April 12^th^ for 2.2% of diagnosed COVID-19 cases in Germany a fatal outcome has been reported [11]. While the epidemic is still ongoing, this formula to calculate the final death rate can be misleading and may lead to a too low number as of today the outcome is unknown for a non-negligible proportion of these diagnosed patients [18]. In other words, current deaths belong to a total case figure of the past, not to the current case figure in which the outcome (recovery or death) of a proportion (the most recent cases) hasn’t yet been determined. Applying the recommended equation to calculate the case fatality ratio (CFR) by death at day D / cases at day D-X and assuming a time of 7 days between reported diagnosis and death an average death rate for Germany of ∼2.9 % can be obtained from the data up to April 12^th^ (total death by April 12^th^ / total cases by April 5^th^) [10].

Assuming 2.5% of diagnosed cases will have a fatal outcome 7 days later we calculated the expected daily deaths in Germany (see Figure 5). The maximum of new deaths per day of ∼150 is expected for April 10^th^. As shown in Figure 5 most of the daily death calculations match well the data. Some differences between the predictions and observed data are evident for the daily new fatalities, which have unexpected high values from April 8 to April 10. This difference may be attributed to COVID-19 outbreaks in nursing homes leading to unexpectedly high case numbers in this period.

It is also noteworthy that with the number of diagnosed COVID-19 cases as input, the assumptions on the length of hospital stays for critical patients and the assumed fraction of 5% critical cases of all diagnosed COVID-19, we expected the maximum of ∼2500 critical cases requiring intensive care with ECMO capability at the first week of April. This is in good agreement with the reported number of COVID-19 cases currently in ICUs in Germany of 2405 on April 12^th^ [1, 11].

In conclusion, we are convinced that regardless of numerous included assumptions derived from clinical observations and regardless of the given uncertainty of the model projections beyond April 10^th^, the model predictions can help to capture the impact of social distancing measures on the epidemic in European countries, to understand the dynamics of the epidemic with respect to diagnosed and fatal cases and to estimate the required daily capacity for German hospitals. Thus, it appears to be a valuable tool for Germany or other countries to provide guidance and support decision making in the health care system with respect to how much hospital capacity is required to be well prepared for the (next) wave.

## Data Availability

For Germany data was obtained from the website of the Robert Koch Institute (RKI) [11]. For Italy the data was taken from the Coronavirus COVID-19 Global Cases published by the Center for Systems Science Engineering (CSSE) at Johns Hopkins University (JHU). For Germany and Italy, we used the reported data up to April 9th for modeling and parameter estimation.

## Acknowledgement

We are thankful to Munkhjargal Schöpfel (Klinikum Koblenz) for sharing her own professional experience with COVID-19 patients, for being a sounding board while this work has been performed and for the supportive daily discussions. We gratefully acknowledge Susana Zaph (Sanofi US) for her thorough and mindful revision of the manuscript.

